# Adjuvant Testosterone Therapy in Chronic Heart Failure (ATTIC) – A Randomised Open-Label Trial

**DOI:** 10.1101/2021.09.07.21262892

**Authors:** Minakshi Dhar, Kartik Mittal, Ashwin Parchani, Manu Sharma, Yogesh Bahurupi, Sanjay Kalra, Nowneet K Bhat

**Author notes:** **Corresponding Author:** Dr. Minakshi Dhar, Additional Professor, Ph No 7060002778, AIIMS Rishikesh. **Author Contributions:** Dr Minakshi Dhar conceptualised the study. Dr Minakshi Dhar and Dr Yogesh Bahurupi designed the study. Dr Kartik Mittal and Dr Ashwin Parchani for the acquisition of data and statistical analyses. Dr Minakshi Dhar, Dr Manu Sharma, Dr Yogesh Bahurupi, Dr Sanjay Kalra and Dr Nowneet K Bhat drafted the manuscript. All authors reviewed the manuscript.

## Abstract

**Introduction:** Heart failure is a major contributor to morbidity and mortality in the geriatric population, with no promising therapy currently available with considerable benefit. Testosterone therapy is an emerging viable treatment option given its beneficial effects, including improving cardiac functional capacity, alleviating symptoms, and low cost, amongst others.

**Methods:** We have planned an open-label, parallel design, 1:1 randomized controlled trial which aims to recruit 160 adult males above the age of 60 diagnosed with chronic stable heart failure fulfilling the eligibility criteria. The participants will be randomized into two groups of 80 each. Both groups will receive standard recommended treatment regime of chronic stable heart failure and intervention arm participants will receive additional testosterone gel. All participants will be assessed at baseline, 4 weeks, 6 weeks and 12 weeks. The primary endpoints will assess the difference in functional capacity, frailty and quality of life at 3 months compared to baseline. The secondary endpoints will include the mean change from baseline at 3 months in cardiac remodelling using echocardiography, serum brain natriuretic peptide levels, the incidence of adverse drug reaction.

**Statistical Analysis:** The data will be analysed with the help of SPSS software. Primary objectives of change in 6MWT, Frailty index and quality of life will be analysed using the student t-test. The statistical significance will be defined as p-value <0.05 and taking confidence level as 95%.

**Ethical clearance:** Institutional Ethics Committee clearance taken via letter no AIIMS/IEC/20/847, dated 21/11/2020

## Introduction

The population share of the elderly is overgrowing worldwide. The number of the elderly is estimated to rise from 524 million in 2010 to nearly 1500 million in 2050, and most of this increase in developing countries.^1^ The factors responsible for this transition are declining fertility, increased longevity and ageing of “baby boom” generations^2^. Heart failure is a unique metabolic syndrome and is rapidly increasing in India due to the rising number of coronary artery disease (CAD), hypertension, diabetes, smoking, alcoholism and the ageing of our society^1^. Due to the unavailability of any promising therapy of heart failure and despite advances in evidence-based pharmacological therapies being evaluated and tested, these patients continue to exhibit significant morbidity and excessive mortality at rates as high as 30% per year^2^, and coupled with ongoing symptoms of fatigue, cardiac cachexia and metabolic shift towards catabolism has led to an intensive search for new therapies Due to this unmet need, novel and low-cost treatments like testosterone hormone replacement therapy are being considered for improving the lives of heart failure patients. Evidence has shown that testosterone improves lean body mass and exercise tolerance and is being investigated worldwide in improving cardiac functional capacity^2^. Chronic heart failure is linked to maladaptive and prolonged neurohormonal and pro-inflammatory cytokine activation, resulting in a metabolic shift favouring catabolism, skeletal muscle atrophy and vasodilator incapacity. Malkin et al. discovered that testosterone replacement treatment improves functional capacity and gives symptomatic relief to men with moderately severe heart failure in a randomized, double-blind, placebo-controlled parallel study of transdermal testosterone therapy^3,4^. In 2014, Mirdamadi et al. investigated the link between low testosterone levels and decreased exercise ability. They aimed to see if testosterone treatment can help individuals with stable chronic heart failure improve their clinical and cardiovascular symptoms and quality of life. In a double-blind, placebo-controlled trial, they randomized 50 male patients with congestive heart failure to receive an intramuscular (gluteal) long-acting androgen injection (1 mL of testosterone enanthate, 250 mg/ml) once every four weeks for 12 weeks or intramuscular injections of saline (1 mL of 0.9 percent w/v NaCl) with the same protocol. The researchers discovered that the two groups’ changes in body weight, hemodynamic parameters, and left ventricular dimensions echocardiographic indices were all comparable. In contrast to the placebo group, those who received testosterone had a substantial increase in 6-wall mean distance (6MWD) during the research (p-value =0.019).^5^.

This study is aimed to improve the quality of life in chronic heart failure patients in the Indian subpopulation of older men, so that transdermal supplementation of testosterone can be adopted as an adjunct therapy to the existing treatment protocols.

### Trial design

An open label trial with 1: 1 allocation

### Aim

To Assess the beneficial effect of adjuvant testosterone therapy in older men with chronic stable Congestive heart failure.

### Primary Objectives

- To determine the effect of testosterone replacement therapy on the functional capacity of older men with chronic congestive heart failure.
- To determine the effect of testosterone replacement on the frailty of older men with chronic congestive heart failure.
- To assess the effect of testosterone on metabolic parameters of older men with chronic congestive heart failure.

### Secondary Objectives

- To assess disease progression by evaluating the status of cardiac remodelling in older men with chronic congestive heart failure by electrocardiography.
- To determine the effect of transdermal testosterone therapy on Brain natriuretic peptide levels in older men with chronic congestive heart failure.
- To study the safety profile of transdermal testosterone therapy in chronic congestive heart failure.

## Material and Method

### Study design

This study will be a randomized open-label clinical trial, parallel design with a 1:1 allocation ratio

### Study Setting for recruitment

The study will be conducted in outpatient and inpatient of the geriatric patient, department of internal medicine in tertiary care level teaching hospital

### Study Duration

The study will be conducted over two years

## ELIGIBILITY CRITERIA

### 1. Inclusion Criteria

Male participants, ≥ 60 years of age, in outpatient and inpatients, who have-

- Chronic stable heart failure with NYHA class II or III irrespective of etiology and whose treatment has not been modified in the past four weeks.
- Testosterone treatment-naive patients.
- Testosterone level low or in the lower half of normal range.
- Patients are willing to provide written informed consent.

### 2. Exclusion criteria

- Any contraindication to transdermal testosterone therapy.
- Deranged Prostate-specific antigen profile (cut off-4 ng/ml)
- Deranged liver function.
- Deranged renal function (serum creatinine level ≥ 1.5 mg/dL)
- Malignancy.

#### Sample size

**t-tests -** Means: Difference between two independent means (two groups)

**Analysis: A priori;** Compute required sample input:

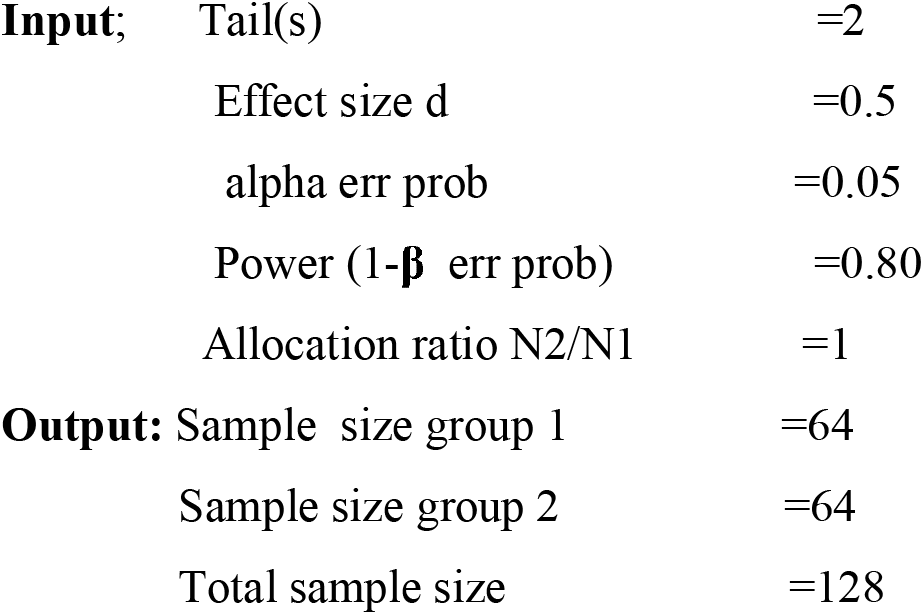

The sample size was estimated after assuming Cohen’s effect size convention (medium effect size) with G*Power software

We expect a 20% attrition rate; the total sample size would be 160, 80 in each group.

## METHODOLOGY: (in accordance with SPRINT Guidelines 2013, Chan et al)^21^

### Randomisation

Computer generated random allocation sequence will be generated using WINPEPI, etcetera (PEPI-for windows). Block randomization with block size four will be done by one of the YB. SNOSE (sequentially numbered opaque sealed envelopes) will be followed for allocation concealment by MP. KM and MD will do enrolment of the patient and assigning the participants to the intervention.

### Treatment arms

**Treatment arms;**
| **Treatment Arm** | **Drug** |
| --- | --- |
| A | 5mg testosterone gel+ Recommended standard therapy for chronic stable heart failure |
| B | Recommended standard therapy for chronic stable heart failure |

Participants in arm ‘A’ will be receiving 5 gm of testosterone gel on top of optimal medical therapy for 12 weeks. A nurse will dispense the drug. Testosterone is available as a 10-gram pouch. Participants will be told to squeeze half of the pouch on a finger and apply it on the deltoid region by gentle massage over an area equal to the size of one rupee coin. The drug will be withheld if the testosterone level increases to above average.

#### Justification of dose

The low dose of testosterone used in the study is not associated with any significant adverse effects and is postulated to improve hemodynamic characteristics in these patients. Gel preparation is preferred because of its good efficacy, lack of variability and short duration of action (which allows immediate discontinuation in case of adverse events).

#### Choice of comparators

Control will be similar group of patients with heart failure and will be treated based on current standard of care as established in international guidelines.

#### Intervention Modification

Any participant on the trial intervention, who undergoes hospitalisation or death, will be discontinued from the trial, with detailed analysis of the event in each discontinued case at the end of the study

#### Intervention adherence

The participant will be asked by the nurse dispensing the trial therapy to follow up with empty pouches of testosterone gel on the regular pre-defined in-person follow ups.

#### Endpoints

##### Primary endpoints

**At the end of three months**

- Change in Functional capacity assessment by 6-minute walk test (6MWT)
- Change in Frailty assessment by using FRIED criteria of Frailty Phenotype
- Change in Quality-of-life assessment using Kansas City Cardiomyopathy Questionnaire

##### Secondary endpoints

- Status of cardiac remodeling by echocardiography in improved ejection fraction as a prognostic marker for disease progression.
- Change in Status of BNP level as a surrogate marker.
- Adverse drug reactions related to testosterone transdermal patch.

##### Data Collection using performa after written informed consent

evaluation for all the patients will be as follows

1. A performed proforma will be used to collect the patients’ demographic data and examination findings, along with a detailed consent form explaining the study aims, objective, methodology, conflicts and benefits as well as harms of the particular intervention.
2. All patients will undergo an echocardiographic assessment at the beginning and the completion of the study.
3. All patients will undergo baseline laboratory investigations which will be repeated as shown in the timeline table. Testosterone will be done in a fasting state.
4. **Functional capacity** assessment; 6 min walk test (6MWT)^6^ will be performed according to standardized procedures. 6-minute walk test distance of <300 m is a simple and useful prognostic marker of subsequent cardiac death in patients with mild-to-moderate heart failure. 6 MWT is a widely available, accessible, low-cost, and well-tolerated test for the assessment of the functional capacity of patients with HF. Although the cardiopulmonary exercise test (a maximal exercise test) remains the gold standard for the evaluation of exercise capacity in patients with HF, the 6MWT (submaximal exercise test) may provide reliable information about the patient’s daily activity.
4. **Frailty** will be assessed using FRIED criteria of Frailty Phenotype (most widely used and validated frailty assessment tool)^7^. It uses five frailty criteria: weight loss, exhaustion, low physical activity, slowness, and weakness. The sum score of these five criteria classifies people into one of three frailty stages (or groups): not frail (score 0), pre-frail (score 1-2) and frail (score 3-5). Fried frailty phenotype criteria are helpful for geriatric inpatient assessment, despite diagnostic limitations, and has been validated for a large number of clinical settings
5. **Assessment of Quality of life**: This will be done and Kansas City Cardiomyopathy Questionnaire (KCCQ)^8^ comprised of the following clinically relevant domains: physical limitations, symptoms (frequency, severity and change over time, self-efficacy and knowledge, social interference, and health-related quality of life.
6. **Insulin resistance**: Glucose and insulin will be measured after overnight fasting. The blood samples will be collected in 5-ml tubes, immediately placed on ice, and transferred to the biochemistry laboratory, where samples will be processed. A commercially available kit will measure plasma insulin levels by immunoradiometric assay (Siemens ADVIA CENTAUR XP immunoassay system). Insulin resistance will be estimated by the homeostasis model assessment (HOMA-IR).^9^
7. **Echocardiographic Evaluation**: It will be done by 2-dimensional, M mode, and Doppler echocardiogram at baseline and at the end of the study by the same physician unaware of the patient’s clinical details as described previously.^10^

##### Blinding

Open-label trial

##### Timelines

**Timelines:**
| Investigations | Baseline | 4 weeks | 6 weeks | 12 weeks |
| --- | --- | --- | --- | --- |
| CBC | ? |  |  | ? |
| LFT | ? |  | ? | ? |
| KFT | ? |  | ? | ? |
| HBAIC and Blood (F/PP) | ? |  |  | ? |
| Serum insulin levels | ? |  |  | ? |
| Serum Testosterone | ? |  | ? | ? |
| PT/INR | ? |  |  | ? |
| NT pro-BNP | ? |  | ? | ? |
| Chest X Ray | ? |  |  | ? |
| ECG | ? |  | ? | ? |
| Echocardiography | ? |  |  | ? |
| 6MWT Score | ? |  |  | ? |
| Frailty test | ? |  |  | ? |
| KCCQ Score | ? |  |  | ? |
| Adverse drug reaction | ? | ? | ? | ? |

##### Study Flow

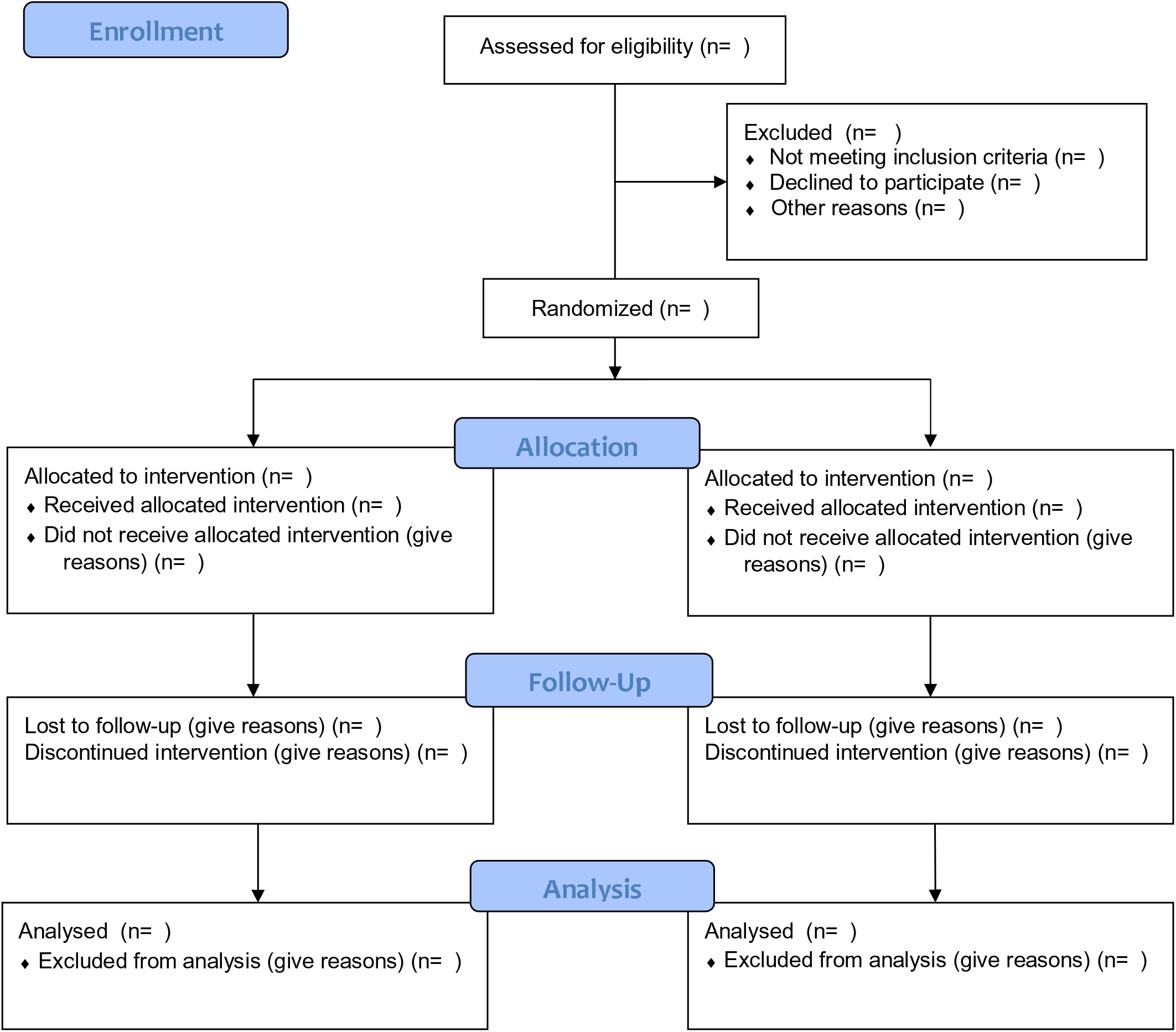

#### Data management

All the data collected during the trial will be stored in an encrypted computer, whose access will be available to the authors of this study only. Due to the unavailability of a data and security management board in our institute, and as the data is going to be used only for academic purposes and there is or role of external industry conflicts of interest, the authors themselves will ensure the data quality is maintained according to best available protocols, which includes collection, storage and analysis, and maintaining a registry of all reported adverse events, including individual details of those cases which have discontinued therapy due to loss of follow up, hospitalisation or death.

### Patient and Public Involvement

Patients of chronic heart failure coming to our OPD will be informed about the study design. They will be informed about study intervention, outcomes both favourable and unfavourable. Written informed consent will be taken from them before recruiting them into the study.

### Statistical Analysis

The data will be tabulated in an excel sheet and analysis will be done with the help of SPSS software. Primary objectives of change in 6MWT, Frailty index and quality of life will be analyzed using the student t-test. Secondary objectives of change in ejection fraction and BNP levels will be done by student t-test, and adverse drug reaction will be represented as percentages. The statistical significance will be defined as p-value <0.05 and taking confidence level as 95%. Data will be analyzed as Per Protocol analysis.

### Confidentiality of data and data access

The data and documents will be kept in confidentiality by the principal investigator. Patients data will not be shared and will be used only for study purposes. Patient identity will be kept confidential. The Institutional Ethics Committee will have access to the data for study-related inspection, if necessary, but only subject to author approval and mentioning of such inspection in final draft of publication.

### Dissemination policy

All the results and analysis, including a detailed discussion of merits, demerits, shortcomings, adverse events and further research will be made accessible to both academicians and public using publication of the data in a research journal, and the access of such a publication will be unrestricted as much as possible, for the use of larger benefit and further studies.

### Ethical clearance

Clearance from the Institutional Ethics Committee of the Institution has been taken via letter no AIIMS/IEC/20/847, dated 21/11/2020. Written informed consent will be obtained from each study participant (Annexure-VI/VII). Information about the study and study procedure will be given with the help of a patient information sheet (PIS) (Annexure-IV/V) in the local language. In the case of the illiterate study participant, contents will be read to the patient in his/her language to their satisfaction by an impartial witness. Any modification in the study protocol will be intimated to Institutional Ethics Committee for approval.

The trial has been registered under the Clinical Trials Registry of India (CTRI). REF/2020/12/030292).

#### Patient Safety

All adverse events detected during the study will be informed to Institute Ethics Committee. The patient’s adverse drug reactions in the intermediate period will be reported to the PI on this mobile number 7060002778. In case of an injury occurring to the participant during the study, free medical management shall be given as long as required, or till such time it is established that the injury is not related to the study intervention, whichever is earlier. In case there is a permanent injury, the quantum of compensation paid shall be as per the clause mentioned in Drugs and Cosmetics Act, 1945.)

## Discussion

This study will be an interventional study in a tertiary care centre in the northern state of India, focusing on heart failure with reduced ejection fraction, using all standard treatment protocols, and adding testosterone, which is approved and easy to use, with better compliance. The project will help in expanding the current knowledge regarding the improvements with current standard treatments and compare them to those where testosterone is added, with data on various parameters where improvement is expected, that include, but are not limited to functional capacity, metabolic parameters and objective cardiac parameters.

Despite the concern about the long-term risk of prostate cancer, currently available data do not support a link between testosterone therapy and prostate cancer. Analysis of 18 prospective studies done by Roddam et al. did not show significant changes in the level of prostate-specific antigen in any individual trial; the pooled estimates indicate a slight, albeit significant, increase in prostate-specific antigen levels.^11^

Low testosterone levels were linked to overall and CV mortality in a meta-analysis by Araujo et al., which comprised of 12 trials with over 17,000 individuals.^12^ Corona et al. included 70 papers in their meta-analysis and showed a clear association between low testosterone and high oestradiol levels and increased CVD and CV mortality risk.^13^ Muraleedharan et al. studied 581 males with Type 2 Diabetes who were followed up for six years. Testosterone treatment was given to 51 males for at least two years. Independent of comorbidities and other therapies, death rates in the low testosterone group was 20.0 percent against 9.1 percent in the normal control, and testosterone therapy decreased mortality equivalent to the controls.^14^ Daka et al. demonstrated that low testosterone concentrations predicted acute MI in men with T2D.^15^ Yeap et al. studied 3,690 older men over ten years and showed that both low and high testosterone levels were associated with all-cause mortality. Ischaemic heart disease mortality was lowered when dihydrotestosterone levels were higher.^16^

A review by Muraldeedharan et al. showed that testosterone level might not be pathological, but low levels may be a marker of illness.^17^

Certain studies pointed out that the use of testosterone can be detrimental to cardiac health, owing to its ability to proliferate myocardial tissue and retain fluids and salt, which can be detrimental in the long run. A study by Basaria et al. in 2012, known as the TOM Trial (Testosterone in Elderly Men with Mobility limitation), showed that in men with mobility limitation, the increase in serum level testosterone can be used to predict the risk of cardiovascular events, wherein those with the highest increase in free serum testosterone level showed the most significant risk of such events, however, logistic regression analysis of this study showed such effect is not significant to owe any harm. Other studies do not support this adverse profiling.^18^ An extensive review of the literature published between 1940 and 2014 found only four studies that reported increased CV risk.^19^

Owing to its proven safety profile, lower cost and wide availability, the use of testosterone has the potential to become a valuable adjuvant to the current standard of care in heart failure, as even the current regimen is not very effective, and all the approved mechanical strategies to relieve heart failure are expensive, albeit not shown to be more effective than conventional treatment modalities.^20^ Owing to this unmet need for a newer, more effective intervention, our study aims to fill this lacuna by including improvement not just in objective parameters but also symptom-based questionnaires to, in a way, some extent proves benefit in quality of life in such elderly patient, which in recent past has become our prime aim as physicians caring for the increasing population of elderly.

## Data Availability

All Data will be made available

## Acknowledgement

We acknowledge our Institute for proving us with the funds and other logistic support to carry out this research. This research is not industry sponsored.

